# Plasma IL-6 Levels following Corticosteroid Therapy as an Indicator of ICU Length of Stay in Critically ill COVID-19 Patients

**DOI:** 10.1101/2020.07.02.20144733

**Authors:** Samir Awasthi, Tyler Wagner, AJ Venkatakrishnan, Arjun Puranik, Matthew Hurchik, Vineet Agarwal, Ian Conrad, Christian Kirkup, Raman Arunachalam, John O’Horo, Walter Kremers, Rahul Kashyap, William Morice, John Halamka, Amy W. Williams, William A. Faubion, Andrew D. Badley, Gregory J. Gores, Venky Soundararajan

## Abstract

Intensive Care Unit (ICU) admissions and mortality in severe COVID-19 patients are driven by “cytokine storms” and acute respiratory distress syndrome (ARDS). Interim clinical trial results suggest that the corticosteroid dexamethasone displays superior 28-day survival in severe COVID-19 patients requiring ventilation or oxygen. Among 16 patients with plasma IL-6 measurement post-corticosteroid administration, a higher proportion of patients with an IL-6 value over 10 pg/mL have worse outcomes (i.e. ICU Length of Stay > 15 days or death) when compared to 41 patients treated with non-corticosteroid drugs including antivirals, tocilizumab, azithromycin, and hydroxychloroquine (p-value = 0.0024). Given this unexpected clinical association between post-corticosteroid IL-6 levels and COVID-19 severity, we hypothesized that the Glucocorticoid Receptor (GR or NR3C1) may be coupled to IL-6 expression in specific cell types that govern cytokine release syndrome (CRS). Examining single cell RNA-seq data from bronchoalveolar lavage fluid of severe COVID-19 patients and nearly 2 million human cells from a pan-tissue scan shows that alveolar macrophages, smooth muscle cells, and endothelial cells co-express both NR3C1 and IL-6. The mechanism of Glucocorticoid Receptor (GR) agonists mitigating pulmonary and multi-organ inflammation in some COVID-19 patients with respiratory failure, may be in part due to their successful antagonism of IL-6 production within lung macrophages and vasculature.

## Introduction

Infection with SARS-CoV-2 is largely asymptomatic or presents with mild-to-moderate symptoms in a majority of patients^1^, but can result in progressive respiratory illness leading to acute respiratory distress syndrome (ARDS) in a subset of severely ill COVID-19 patients^2,3^. Multiple clinical trials are underway to investigate therapies ranging from antivirals (e.g. Lopinavir-Remdesivir) to corticosteroids and immunosuppressive agents (e.g. Dexamethasone, Tocilizumab)^4^. In line with previous reports of IL-6 as a biomarker of severe disease^5^, some studies examining small patient cohorts have suggested treatment with tocilizumab (anti-IL6 receptor) may improve outcomes in severe COVID-19 patients^6,7^. Consequently, the use of plasma IL-6 testing has gained traction during the ongoing COVID-19 pandemic and suggests IL-6 as a promising biomarker to study in the context of COVID-19.

We were motivated by the observation that high sensitivity plasma IL-6 tests conducted at the Mayo Clinic between 2010 and 2019 show only 274 out of 1463 tests (18%) above 10 pg/mL (normal range between 0.31-5 pg/mL), whereas 377 out of 537 tests (70%) in the first half of 2020 alone are above 10 pg/mL (**Figure 1 - Supplementary Figure 1**). The distribution of observed IL-6 levels during the ongoing COVID-19 pandemic (mean = 68 pg/mL) is thus shifted towards higher levels compared to the IL-6 levels from earlier years (mean = 11 pg/mL). We also observed that IL-6 levels did not correlate with levels of the inflammatory markers C-reactive protein (CRP) and erythrocyte sedimentation rate (ESR) in the COVID_pos_ patients (**Figure 1 - Supplementary Figure 2**). Spurred by these observations, we were keen to analyze the retrospective real-world evidence regarding the use of plasma IL-6 levels to study the prognosis of severe COVID-19 patients. We were also intrigued by the availability of a small cohort of patients with serial measurements of plasma IL-6 levels tied to the therapeutic regimen administered and clinical outcomes recorded in the associated EHRs.

A recent interim update from the RECOVERY (Randomised Evaluation of COVid-19 thERapY) trial that is examining larger cohorts of severe COVID-19 patient outcomes revealed the maximal reduction in mortality among patients treated with dexamethasone over other therapies^8^. In this trial, 2,104 randomized patients received 6 mg of dexamethasone once per day for ten days, compared with 4,321 patients that received standard care alone. Dexamethasone reduced mortality by one-third in ventilated patients (rate ratio 0.65 [95% confidence interval 0.48 to 0.88]; p=0.0003) and by one-fifth in other patients receiving oxygen only (0.80 [0.67 to 0.96]; p=0.0021)^8^.

Although corticosteroids have long been utilized clinically for their immunosuppressive and anti-inflammatory capacities^9,10,11,12^ the precise mechanisms by which dexamethasone mediates clinical improvement in severe COVID-19 patients are not well understood. Indeed, the success of dexamethasone in treating COVID-19 was somewhat unexpected in light of an early report during this pandemic urging clinicians to not prescribe corticosteroids for lung injury in COVID-19 patients, based on the lack of efficacy as well as elevated risk of adverse events associated with steroid use in the previous SARS and MERS epidemics^13^. Such ostensibly contradictory evidence and guidance underline the need for an improved mechanistic stratification of corticosteroid efficacy in different subsets of severely ill COVID-19 patients.

Rapid advances in genomic and transcriptomic technologies over the past decade hold great potential to characterize drug targets at unprecedented levels. We recently released the n*f*erX platform Single Cell application as a resource to help researchers analyze publicly deposited single cell RNA-sequencing datasets and readily contextualize these expression-derived insights using quantified literature associations^14^. We and others have harnessed this wealth of gene expression data at single cell resolution to profile human tissues and cells based on their expression of ACE2, the putative entry receptor for SARS-CoV-2^14,15,16,17^. Notably, although the primary glucocorticoid receptor (NR3C1) has been previously reported to be ubiquitously expressed^18,19,20^, the global expression profile of this important drug target has not been systematically evaluated across the hundreds of thousands of bulk RNA-seq samples and millions of single cell RNA-sequencing data points which are available.

Here, we perform longitudinal analysis of real-world data and COVID-19 patients and triangulate our findings with publicly available transcriptomic datasets to nominate putative mechanisms by which dexamethasone results in reduced mortality. Our findings provide molecular support for the known immunomodulatory effects of corticosteroids and pinpoint the immune cell types which are most likely to be affected by systemic and pulmonary exposure to dexamethasone. Specifically in one study of COVID-19 alveolar immune infiltrates^21^, we identify multiple populations of macrophages which co-express NR3C1 and IL-6 genes, including one coexpressing population which tends to downregulate NR3C1 in severely ill patients compared to those with mild disease. We also find that non-immune cells including endothelial cells and smooth muscle cells are among the cell types which most frequently coexpress these genes.

Taken together, our findings provide initial molecular evidence to suggest that the clinical improvement observed with corticosteroid treatment may be due to NR3C1-mediated antagonism of IL-6 production both systemically and locally in the lungs. Our study warrants detailed follow-up analyses to establish the clinical efficacy of dexamethasone and other corticosteroids in COVID-19, and to identify molecular or cellular biomarkers associated with therapeutic responses.

## Results

### Stratifying COVID-19 patients by ICU duration highlights IL-6 levels post administration of corticosteroids, but not non-corticosteroids, as an indicator of COVID-19 outcomes

To understand whether or not corticosteroids benefited inflammation-induced respiratory distress among the ICU patients, we started by characterizing respiratory status in patients who received corticosteroids and IL-6-testing versus those who received IL-6-testing alone. Among COVID19 patients that were admitted to ICU at some point during their hospital stay and had undergone an IL-6 test, patients that were on corticosteroids experienced longer durations on higher levels of respiratory support compared to those that did not receive corticosteroids (**Figure 1a**).

**Figure 1.**
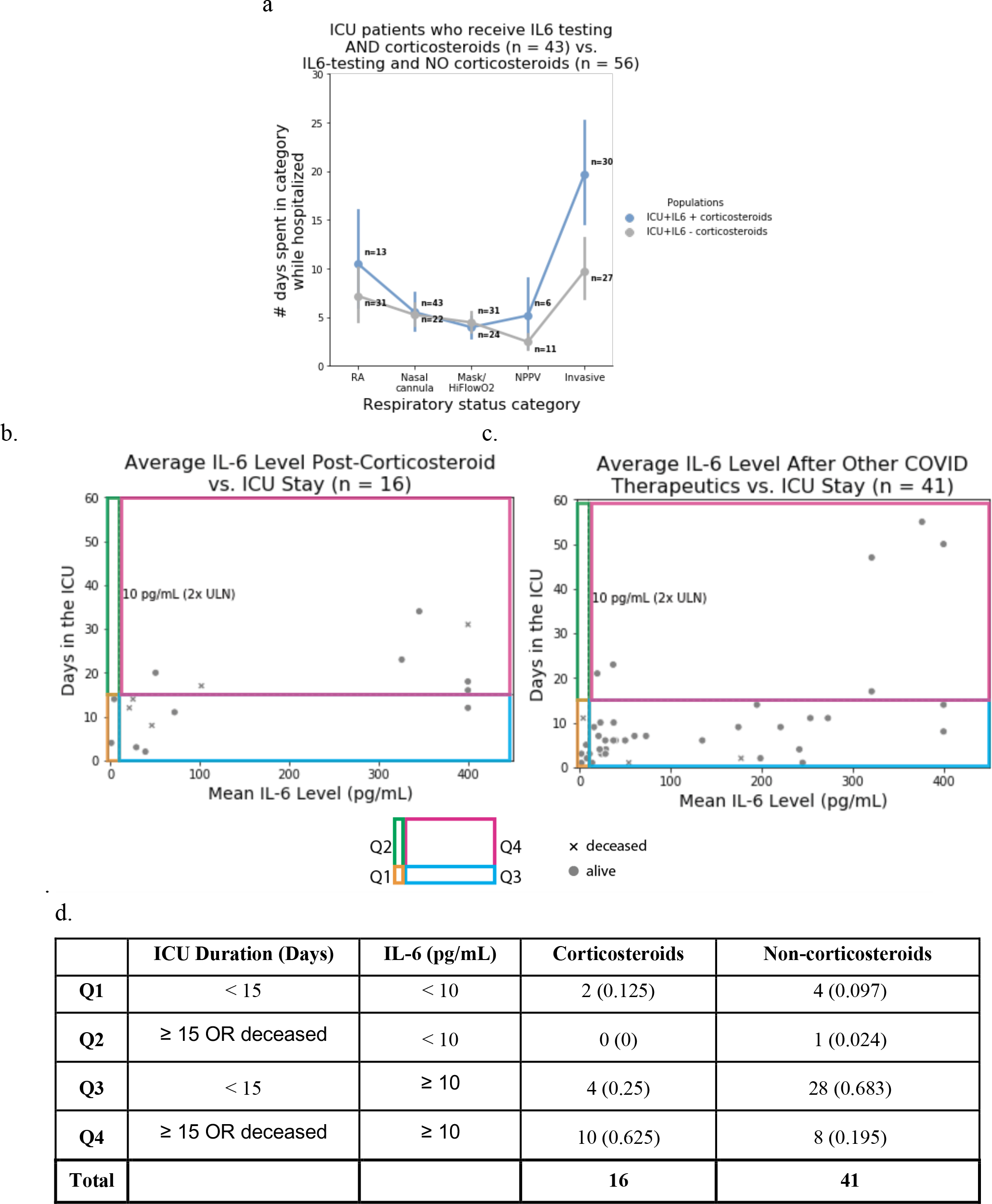
Relationship between IL-6 levels and ICU duration for COVID-19 ICU patients who had at least one plasma IL-6 measurement after administration of either corticosteroids or other classes of drugs. **(a)** Among the COVID19 patients that were admitted to ICU at some point during their hospital stay and had undergone an IL6 test, the patients that were on corticosteroids (n = 43) experienced longer durations on higher levels of respiratory support compared to the patients that were not on corticosteroids (n = 56). (**b**) Relationship between ICU duration and IL-6 levels following the administration of non-topical corticosteroids for n = 21 ICU patients who received corticosteroids. (**c**) Relationship between ICU duration and IL-6 levels following the administration of one or more of the following drugs: Tocilizumab, Azithromycin, Hydroxychloroquine, or antivirals (n = 64 patients); these patients did not receive corticosteroids. **(d)** Summarizing the number of patients in each of the 4 quadrants (Q1, Q2, Q3, Q4) in figures 1b and 1c above. A Fisher exact test of proportions suggests that a higher proportion of patients who receive corticosteroids and have an IL-6 value over 10 pg/mL have poor outcomes (ICU stay >= 15 days or death) when compared to the equivalent proportion of those treated with non-corticosteroid drugs including antivirals, tocilizumab, azithromycin, and hydroxychloroquine (*p = 0*.*0006*).

To understand how corticosteroids either benefited or not the inflammation-induced respiratory distress among the ICU patients, we plotted plasma IL-6 levels versus ICU durations for patients on corticosteroid and non-corticosteroid therapies (antivirals, azithromycin, hydroxychloroquine, tocilizumab). Thresholds were set for IL-6 level (10 pg/mL) and ICU status (15 days) to group patients into four categories, with mortality grouped with ICU stay of 15 days or more. Among 16 patients with plasma IL-6 measurement post-corticosteroid administration (**Figure 1b**), a higher proportion of patients with an IL-6 value over 10 pg/mL have worse outcomes (i.e. ICU stay > 15 days or death) when compared to 41 patients treated with non-corticosteroid drugs (**Figure 1c**). The Fisher’s exact test p-value for this observation is 0.0024 (**Figure 1d**). The split-up of non-corticosteroid drugs into the specific therapeutics is also shown (**Figure 1 - Supplementary Figure 3**), with consistent observations to those summarized above.

### Longitudinal comparison of plasma IL-6 levels pre- versus post-administration of corticosteroids

The timing of SARS-CoV-2 PCR testing, non-topical corticosteroid administration, tocilizumab administration, and IL-6 measurements available for 63 hospitalized COVID-19 patients are summarized in **Figure 2** - **Supplementary Figure 1**. Of these patients, we considered 10 critically ill COVID-19 patients who received IL-6 plasma tests both before and after corticosteroid administration. These patients met the following criteria: (1) hospitalized in the ICU for COVID-19 related illness, (2) received non-topical corticosteroid therapy during COVID-19 related hospitalization, and (3) underwent plasma IL-6 testing at least one time both before and after the first administration of a corticosteroid. Of these patients, 4/10 were male and the median age was 62 (interquartile range: 55-69).

We found that IL-6 levels remained stable or decreased after steroid administration in 7 of 10 patients, while the remaining 3 patients showed large increases in IL-6. (**Figure 2a**). Nine of the 10 patients began with pre-corticosteroid IL-6 levels more than twice the upper limit of normal (normal reference range: 0.31-5 pg/mL). Following corticosteroid administration, 5 of the 10 patients returned to mean IL-6 levels less than twice the upper limit of normal (10 pg/mL) and 4 of the 10 patients had IL-6 levels less than 5 pg/mL. The was a weak correlation between change in average plasma IL-6 levels following corticosteroid administration and worse clinical outcomes (**Figures 2b**). However, an ICU duration of 15 days or more was not significantly associated with post-steroid increases in IL-6 (p-value = 0.17; **Figure 2c**). Thus, longitudinal IL-6 testing may be a useful biomarker for COVID-19 severity and resolution, but larger patient numbers would be required to confirm this hypothesis.

**Figure 2.**
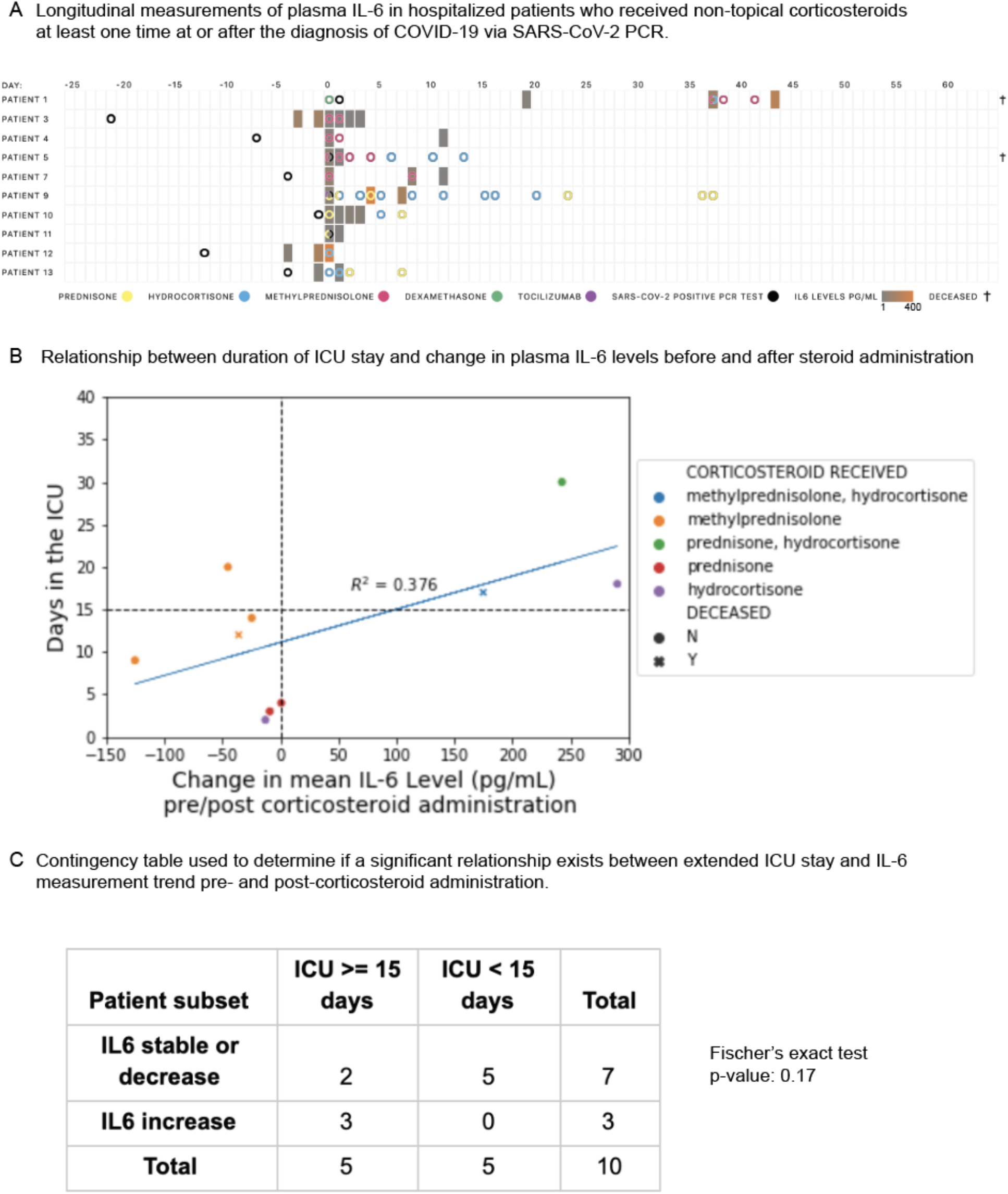
Constraining cohort of COVID-19 ICU patients to those who had at least one plasma IL-6 measurement before and after corticosteroid administration. (**A)** Longitudinal measurements of plasma IL-6 in hospitalized patients who received non-topical corticosteroids at least one time at or after the diagnosis of COVID-19 via SARS-CoV-2 PCR. **(B)** Relationship between duration of ICU stay and change in plasma IL-6 levels before and after steroid administration (patient numbers are mapped onto Figure 8a). If multiple pre/post-corticosteroid administration levels are available, the mean is used. (**C**) Contingency table used to determine if a significant relationship exists between extended ICU stay and IL-6 measurement trend pre- and post-corticosteroid administration.

### The corticosteroid target NR3C1 is strongly expressed across hematopoietic cell types including in alveolar macrophages

To better understand the mechanisms of potential corticosteroid-induced effects on IL-6 levels, we next systematically profiled the gene expression of NR3C1 - the highest affinity target of dexamethasone, methylprednisolone, and prednisone (**Figure 3 - Supplementary Figure 1**) - across over 450,000 human bulk RNA-sequencing samples from more than 10,000 studies. Such an analysis is timely and critical to identify potential mechanisms of steroid-induced immunomodulation. Indeed, the immunomodulatory function of corticosteroids has long been appreciated^9,10^, yet the expression profile of NR3C1 has not been formally established across all human tissues and cell types. The delineation of this profile in both healthy and diseased patients may help us to better understand the cell types which are most critical to mediate the beneficial effects of steroids in severely ill COVID-19 patients. We found that various immune cells - including T cells, B cells, monocytes, natural killer cells, dendritic cells, and macrophages - are enriched among the samples with the highest (top 5%) of NR3C1 expression (**Figure 3a**), consistently with previously reported effects of glucocorticoids on the function of each of these cell types^23,24,25,26,22^. Interestingly, we also identified one study in which alveolar macrophages specifically showed high NR3C1 expression (**Figure 3 - Supplementary Figure 2;** n = 20 samples from 10 individuals), and we found that NR3C1 was consistently expressed at appreciable levels in alveolar macrophages from two other independent studies as well (**Figure 3 - Supplementary Figure 2**; n = 25 samples).

**Figure 3.**
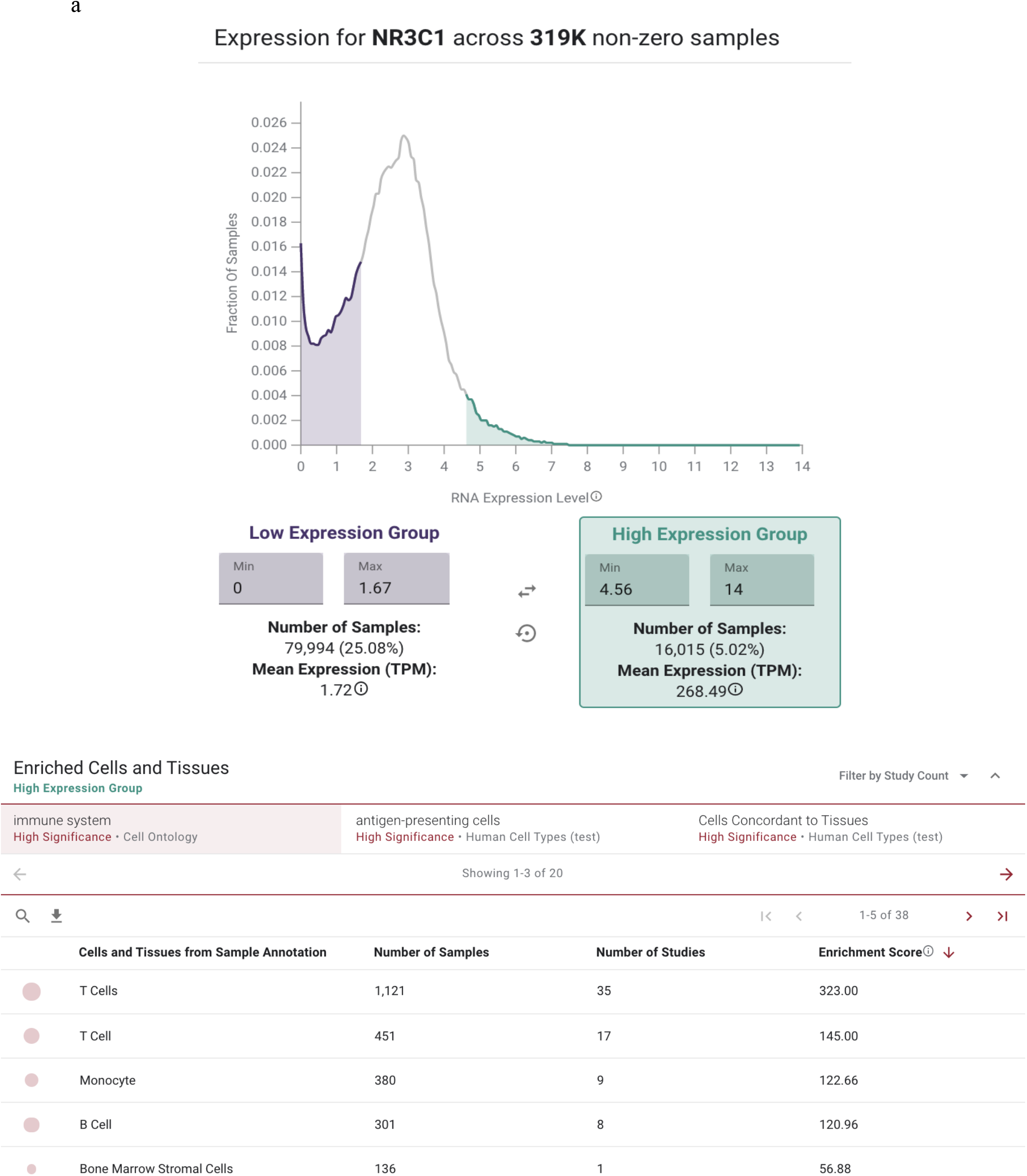
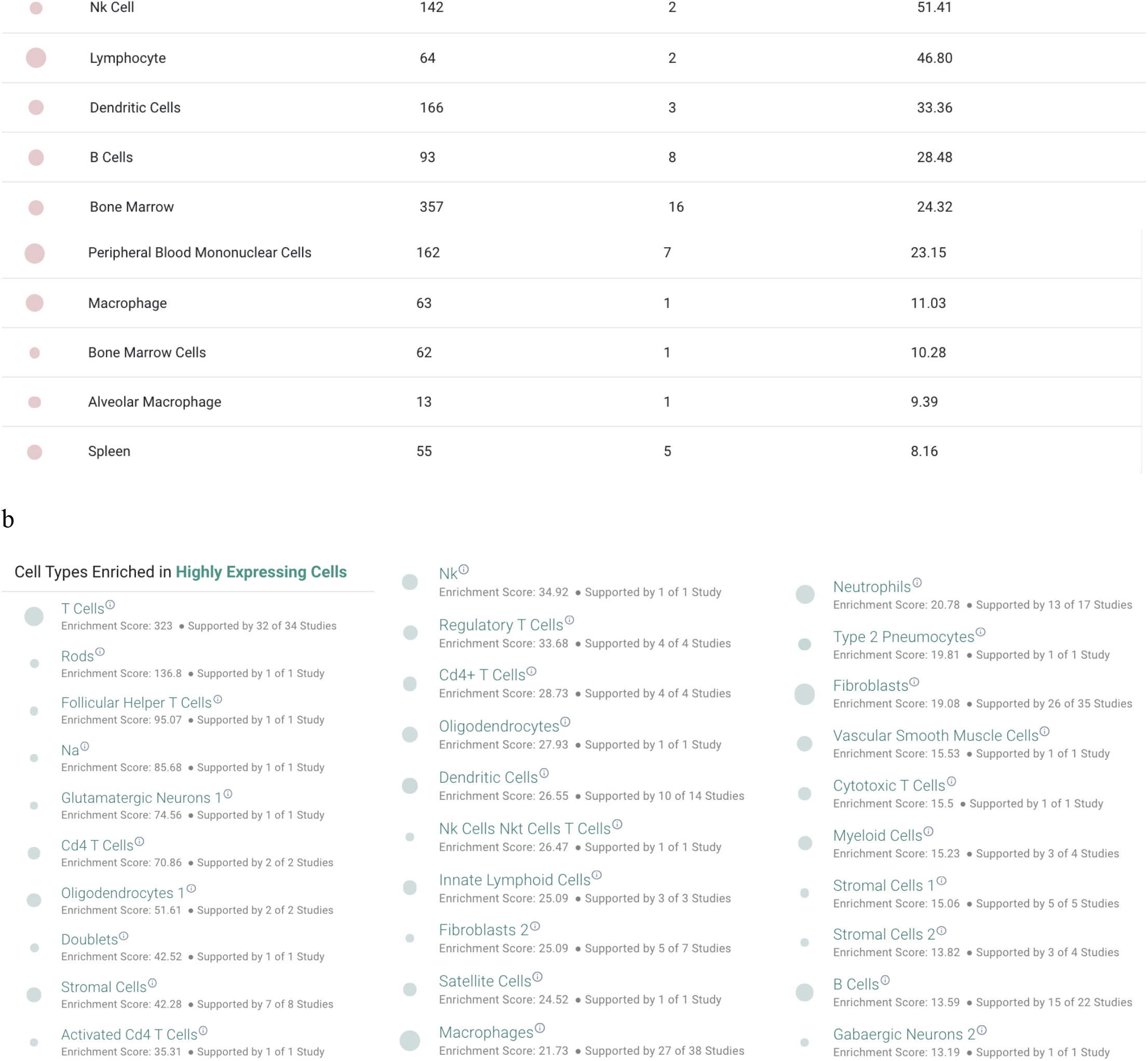
(**A**) Expression profile of NR3C1 across approximately 319,000 non-zero expressing bulk RNA-sequencing samples from the Gene Expression Omnibus. Cells of the immune system are enriched among the highest expressing samples, including both lymphocytes and myeloid cells. (**B**) Single cell RNA-sequencing expression profile of NR3C1 across approximately 1.9 million individual human-derived cells. Cells of the immune system are enriched among the highest expressing samples, including both lymphocytes and myeloid cells.

We then further characterized NR3C1 expression by single cell RNA-sequencing across 1.9 million human cells using the nferX Single Cell platform^14^. Consistent with our findings from bulk RNA-seq, we found that various hematopoietic lineages were enriched among the cells with highest expression of NR3C1 (**Figure 3b**). Several T cell populations showed particularly strong enrichment among NR3C1-high samples, but we also found evidence for high expression in macrophages, dendritic cells, B cells, and innate lymphoid cells (**Figure 3b**). Similarly, in a study of peripheral blood samples from healthy donors (n = 6) and COVID-19 patients between days 2 and 16 of symptom onset (n = 7), NR3C1 was appreciably expressed in diverse immune cell types including T cells, B cells, dendritic cells, monocytes, and plasmacytoid dendritic cells (**Figure 4**). In bronchoalveolar lavage fluid (BALF) of COVID-19 patients, both macrophages and T cells strongly expressed NR3C1, while plasma cells, neutrophils, and the recovered epithelial populations showed lower levels of NR3C1 expression (**Figure 5**; <u>nferX platform Single Cell app</u>).

**Figure 4.**
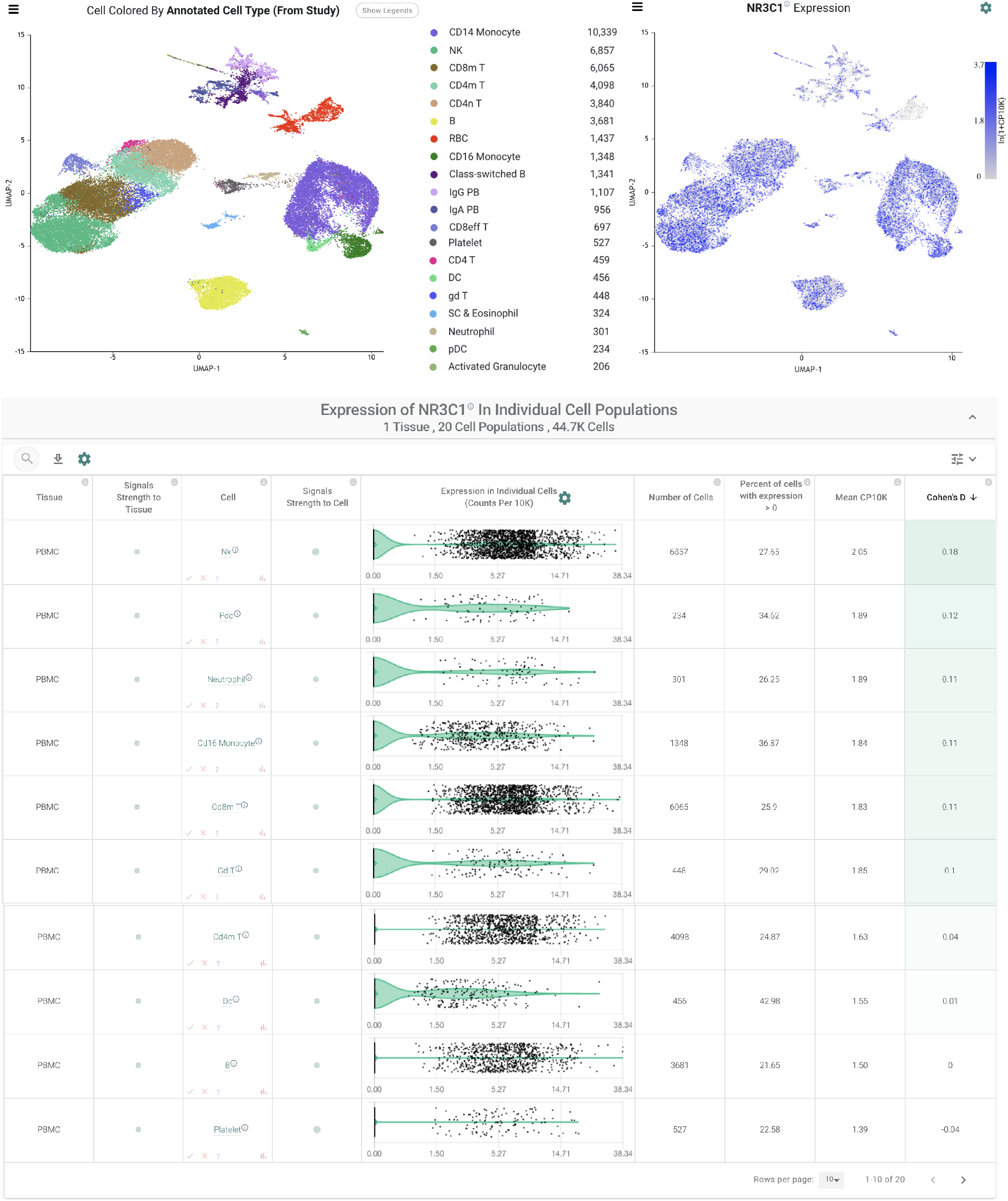
Expression of NR3C1 in peripheral blood mononuclear cell populations from healthy donors (n = 6) COVID-19 patients (n = 7) by single cell RNA-sequencing.

**Figure 5.**
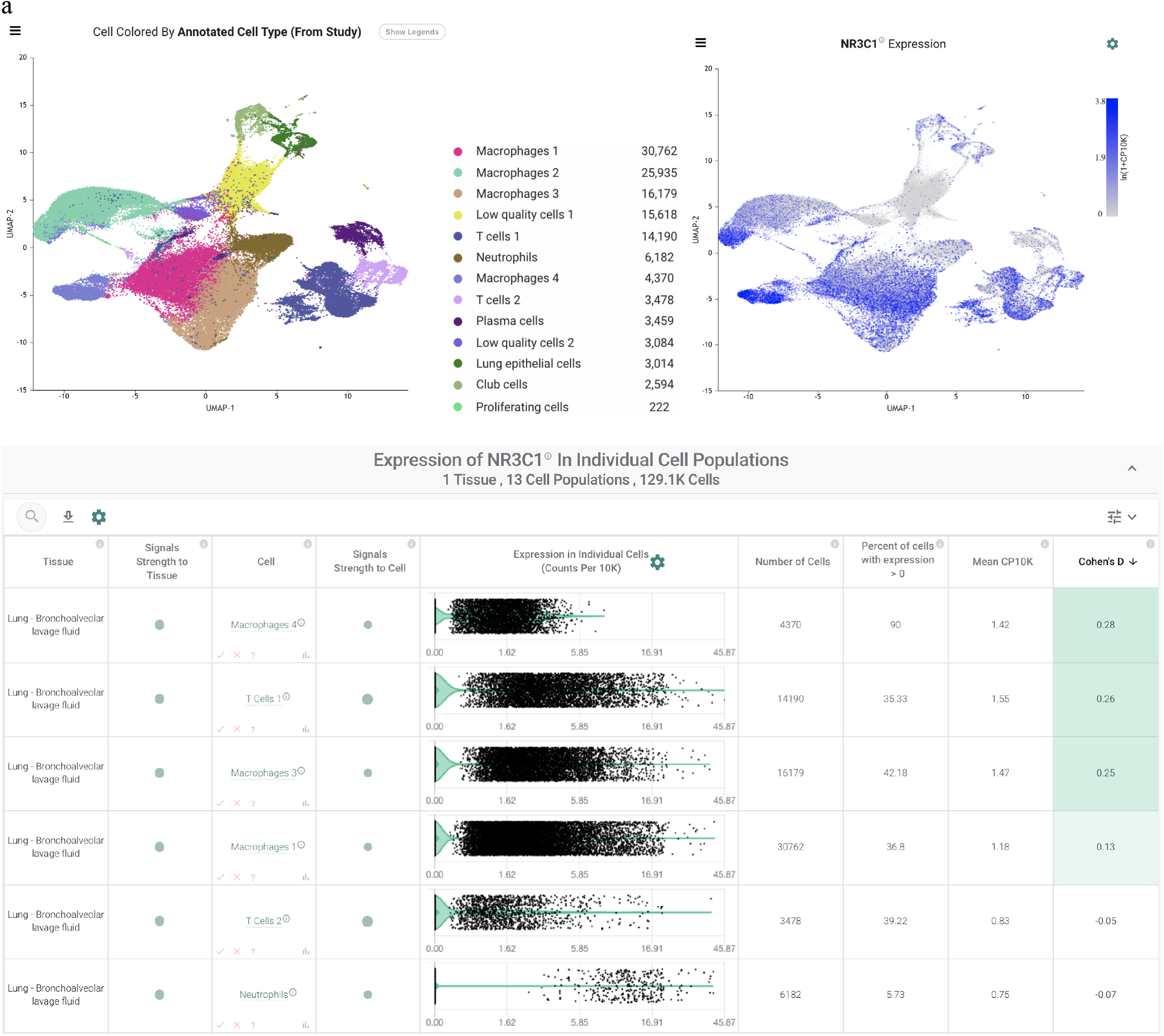
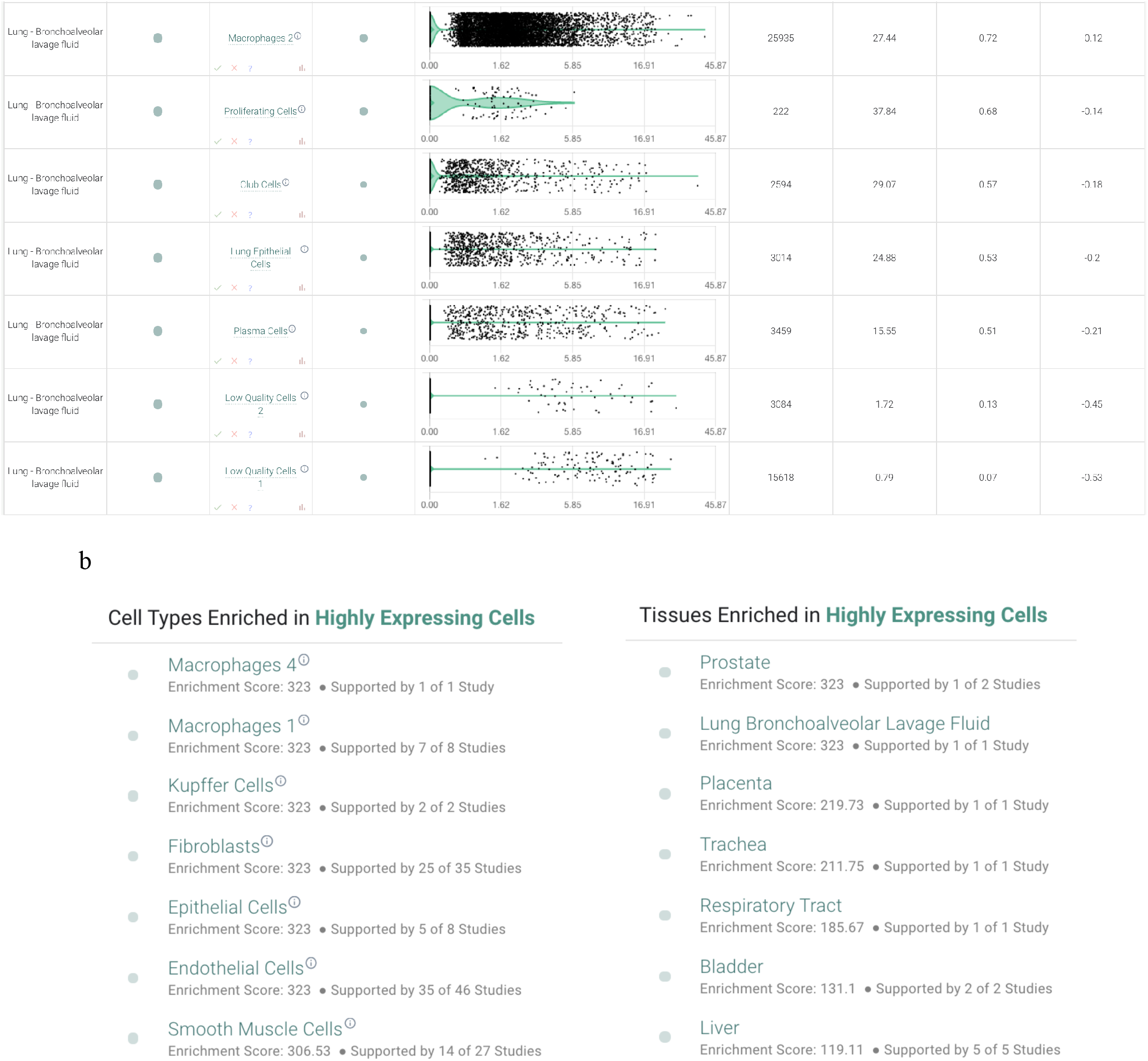
(**A)** Expression of NR3C1 in cells from bronchoalveolar lavage fluid (BALF) of COVID-19 patients (n = 9; 3 mild, 6 severe) by single cell RNA-sequencing. (**B**) Summary of IL-6/NR3C1 co-expression across 1.9 million human cells from single cell RNA-sequencing datasets. Shown are cell and tissue types most highly enriched for detection of both NR3C1 and IL-6 in the same individual cells.

### NR3C1 and IL-6 are co-expressed in alveolar macrophages of COVID-19 patients and systemically in endothelial cells and smooth muscle cells

To connect this expression profile to our clinical observation of reduced plasma IL-6 in a subset of patients following corticosteroid administration, we specifically evaluated co-expression of NR3C1 and IL-6 across these same 1.9 million human single cells. We found that two populations of alveolar macrophages from a study of healthy controls (n = 3) and COVID-19 patients (n = 9) were among the most strongly enriched cell types for this co-expression, with both genes detected in 20% and 5% of these two populations (“Macrophages 4” and “Macrophages 1”, respectively; **Figure 6a**). Interestingly, in contrast to the immune-centric expression profile of NR3C1 alone, we found that several other non-immune cell types showed notable co-expression with IL6 in both respiratory (**Figure 6b**) and non-respiratory (**Figure 6c-d**) tissues including fibroblasts, epithelial cells, endothelial cells, and smooth muscle cells. This observation of co-expression in both endothelial cells and smooth muscle cells is particularly interesting given the reports of systemic vascular inflammation in the context of COVID-19^27–29^. Given the well-established mechanisms of glucocorticoid-mediated suppression of IL-6 transcription^30^, we propose that agonism of NR3C1 in these various co-expressing cell types serves to dampen IL-6 production both in the lungs and systemically.

**Figure 6.**
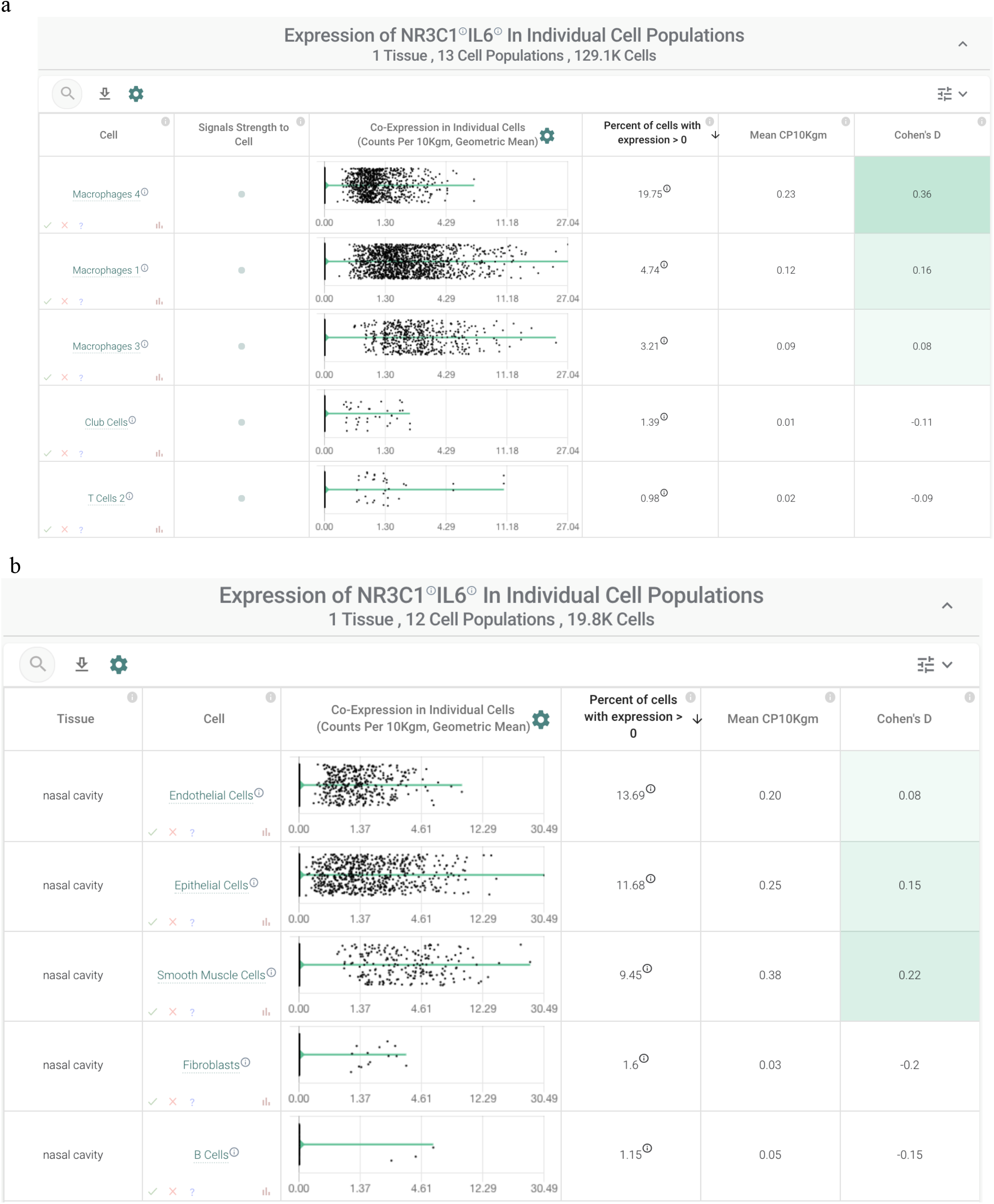
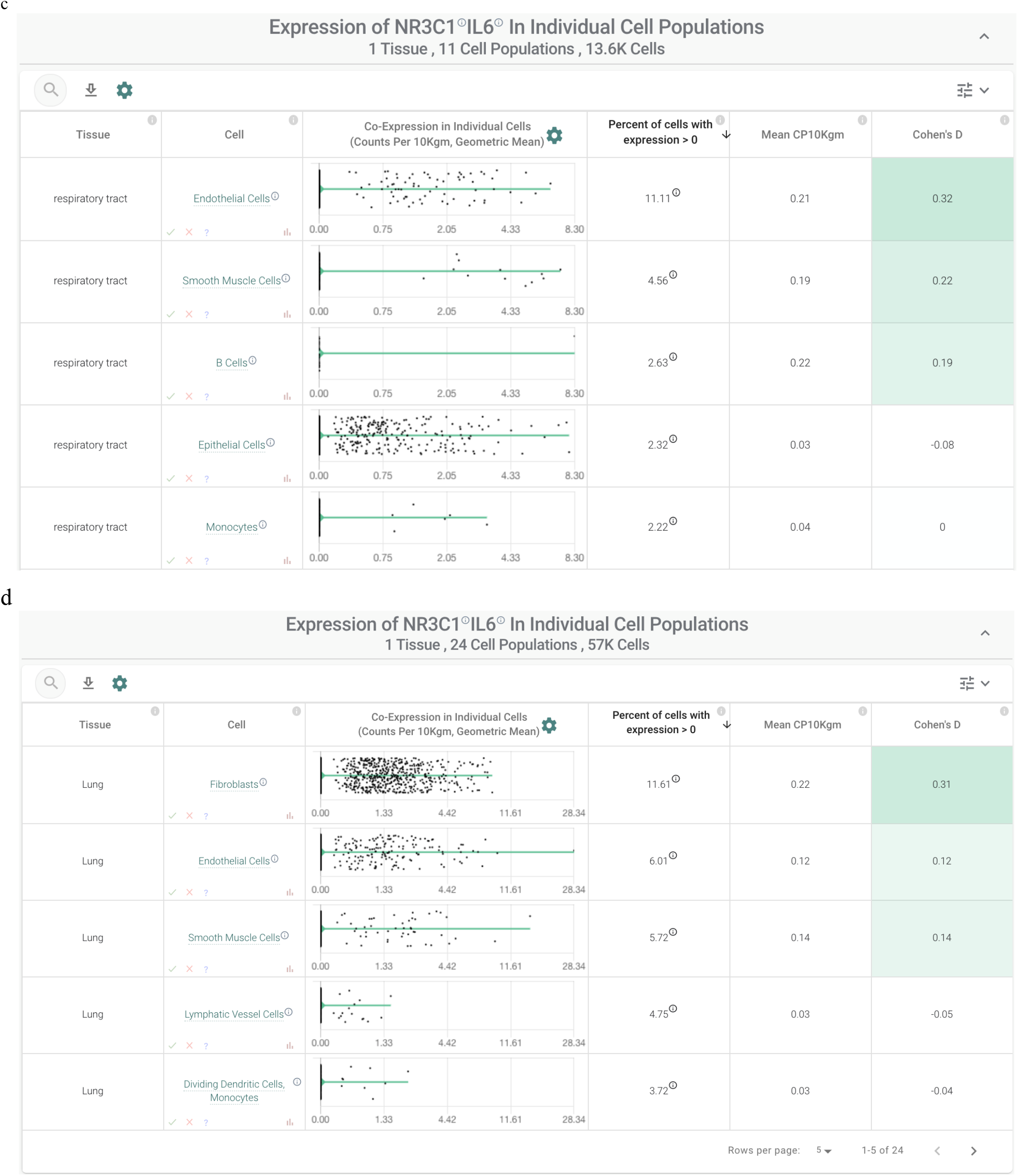
IL-6/NR3C1 co-expression by single cell RNA-sequencing in cells from: (**A**) bronchoalveolar lavage fluid of COVID-19 patients; and **(B-D)** nasal cavity, respiratory tract, lungs.

### In severe COVID-19 patients, NR3C1 is downregulated in macrophages that co-express IL-6 and NR3C1

Given that dexamethasone appears to reduce mortality among only severe cases of COVID-19, we finally asked how NR3C1 and IL-6 co-expression varies between mild and severe disease. To answer this, we assessed NR3C1 and IL-6 expression levels in each recovered bronchoalveolar lavage fluid (BALF) cell population between cases of mild COVID-19 (n = 3) and severe COVID-19 (n = 6), along with healthy controls (n = 3). Interestingly we found that expression of IL-6 and NR3C1 is positively correlated in the strongest co-expressing population (“Macrophages 4” identified above), and further that NR3C1 expression in this cell population was significantly lower in patients with severe compared to mild disease (**Figures 7a-b**). We hypothesize that this overall direct correlation may reflect a cell-intrinsic mechanism wherein activated inflammatory macrophages are simultaneously primed for homeostatic steroid-mediated immunosuppression. Indeed, such programmed feedback loops are well-established to dampen immune responses, including most notably the immune checkpoint pathways (e.g. PD-1/PD-L1) which restrain excessive T-cell activation upon antigen recognition^31^. Accordingly, our observation that NR3C1 expression is decreased in severe COVID-19 patients compared to mild COVID-19 patients may reflect a pathologic downregulation of this endogenous immunomodulatory system which can be restored pharmacologically via corticosteroid-mediated agonism of NR3C1. Together with potential broader effects exerted through the various other previously identified NR3C1/IL-6 co-expressing cell types (see **Figure 7**), corticosteroid therapy may thereby dampen both local pulmonary and systemic inflammation to reduce the likelihood of patient progression to outright cytokine storm.

**Figure 7.**
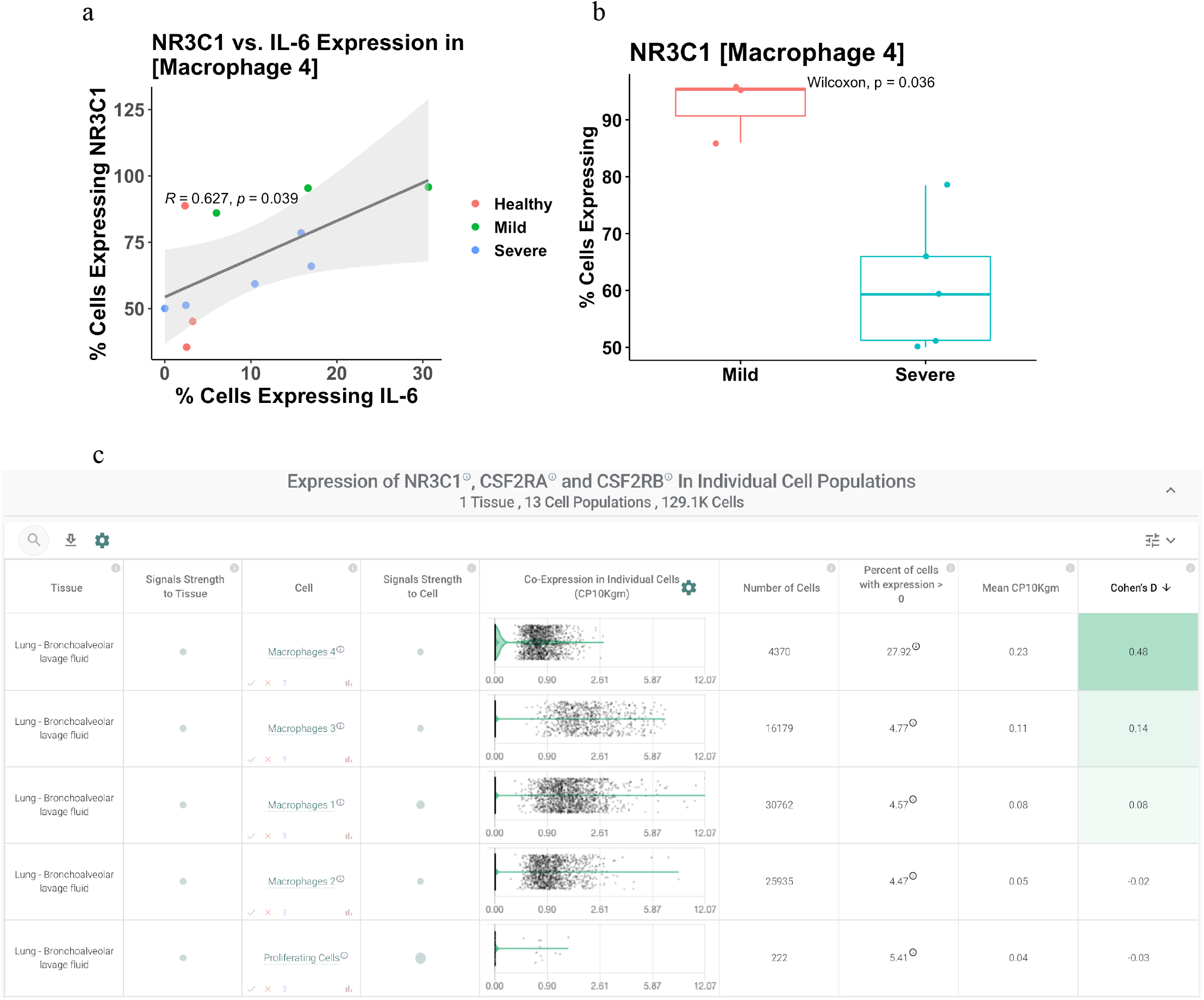
Relationship of NR3C1 and IL-6 expression to COVID-19 status and clinical severity. **(A)** Correlation between IL-6 and NR3C1 expression in the macrophage population found to most robustly co-express these two genes (“Macrophages 4”). Each dot represents the percent of cells from this cluster (i.e. cell population) in which IL-6 and NR3C1 were detected, and dots are colored by COVID-19 status and clinical severity. Values shows are the Pearson correlation coefficient (“R”) and corresponding p-value (“p”). **(B)** Comparison of NR3C1 expression in the “Macrophage 4” cell population from patients with mild versus severe COVID-19. (**C**) Coexpression of NR3C1 with the two subunits of the GM-CSF receptor (CSF2RA and CSF2RB) in the bronchoalveolar lavage fluid of COVID-19 patients (n = 9; 3 mild, 6 severe). Top coexpressing populations, all of which are macrophages, are shown.

## Discussion

The COVID-19 pandemic is unquestionably one of the most urgent global health crises of the modern era. Its onset and dissemination around the world has sparked clinical and basic research in efforts to discover interventions which can effectively eliminate infection or mitigate the clinical course of disease progression. While antiviral drugs, antibiotics, and antimalarial have shown little to no clinical benefit in COVID-19 patients^32,33,34^, modulation of host inflammatory responses with drugs that inhibit IL-6 signaling (tocilizumab) or stimulate glucocorticoid receptor activity (dexamethasone) have been shown to improve clinical outcomes in critically ill patients^6,35^. This is consistent with the recent establishment of IL-6 as a biomarker of COVID-19 severity^5^.

Along these lines, a summary of the ICD-10 codes enriched among patients with high plasma IL-6 levels in 2019 included “Nonspecific Abnormal Finding of the Lung” whereas those enriched in 2020 include “Acute Respiratory Failure with Hypoxia” and “COVID-19 Infection”. This highlights the strong clinical association between high IL-6 levels and severe COVID-19 infection, and motivates better understanding of whether the suggested clinical benefit of corticosteroid therapy in critically ill COVID-19 patients may be related to their established ability to reduce the transcriptional expression of IL-6 and other inflammatory mediators in various cell types^22^.

In this study, we performed an integrated analysis of publicly available molecular data and curated EHR data as well as associated lab test results to study mechanisms of corticosteroid mitigation of COVID-19 cytokine storms and ARDS. Strikingly, despite the long history of corticosteroid use in the clinic, expression of the glucocorticoid receptor (NR3C1) has never been systematically profiled at single-cell resolution in the modern genomic era. Such a profiling will help understand the tissues and cell types which are most likely to be directly targeted by systemic steroid therapies. Consistent with the well-established and clinically utilized anti-inflammatory effects of corticosteroids, our analysis of gene expression by bulk and single cell RNA-sequencing strongly suggests that hematopoietic cells are the cells most prominently impacted by the corticosteroid dexamethasone via its NR3C1 agonism. Specifically, we find that T cells are the most strongly enriched human cell type for high NR3C1 expression, but other adaptive and innate immune cells are also notably strong expressors.

After NR3C1, the second highest affinity target known for the corticosteroid dexamethasone is NR3C2, which was not explored in this study. Primarily, the intersection between IL6 and NR3C2 appears to be limited to the ovaries (**Figure 3 - Supplementary Figure 3**). The NR3C2 activity of corticosteroids, and indeed the other targets including progesterone receptors, may well be beneficial for other aspects of COVID-19 severity mitigation beyond the IL-6 cytokine nexus that was explored in this study via clinical-omics triangulation. The polypharmacology of corticosteroids as they pertain to COVID-19 will be the topic of a follow-up study.

To understand the cellular context around the intersection of glucocorticoids and IL-6, in this study, we have performed the first directed co-expression analysis of NR3C1 and IL-6 across human single cell RNA-sequencing datasets totaling almost 2 million cells. Interestingly, this analysis highlighted that while NR3C1 alone is highly expressed in T cells throughout the human body, co-expression with IL-6 is prominently observed in alveolar macrophages of the lung along with various non-immune cells across multiple tissues including endothelial, smooth muscle, epithelial, and stromal cells. We hypothesize that the engagement of the glucocorticoid receptor by dexamethasone in these co-expressing cell types reduces local and systemic IL-6 production, which in turn restores immune homeostasis and mitigates the progression of the COVID-19 associated acute respiratory distress syndrome. In support of this hypothesis, our analysis of EHR data from a large academic medical center shows that plasma IL-6 levels are reduced after steroid administration in a subset of critically ill COVID-19 patients, and patients with this steroid-associated IL-6 decline tend to have shorter ICU admissions than patients whose IL-6 levels increase even after steroid administration.

It is important to note that other inflammatory pathways are also notably activated - and excessively so - in COVID-19 patients and that blockade of these pathways may provide clinical benefit similar to that seen with tocilizumab. For example, early results indicate that blockade of the GM-CSF pathway with the monoclonal antibody mavrilimumab may improve outcomes in several ill patients. GM-CSF is known to orchestrate the activity of various innate immune cells including dendritic cells^36^ and macrophages^36^, and single cell RNA-seq data confirms that NR3C1 is co-expressed with both the alpha and beta subunits of the GM-CSF receptor (CSF2RA and CSF2Rb) in several macrophage populations from the BALF of COVID-19 patients (**Figure 7c**). Whether agonism of the glucocorticoid signaling pathway in these cells impacts their response to GM-CSF is certainly relevant to investigate as this could have direct clinical implications regarding the utility of coadministration of corticosteroids with both tocilizumab and mavrilimumab.

The results reported in this study emphasize that follow-up studies with larger cohorts of longitudinal data are warranted to investigate the clinical efficacy of dexamethasone and other corticosteroids in COVID-19 patients with ARDS and CRS. Such larger follow-up studies will help shed light on the mechanisms underlying heterogeneity in patient responsiveness to corticosteroids. If suppression of IL-6 production by corticosteroids like dexamethasone is indeed confirmed to be responsible for mitigating the respiratory and vascular onslaught of SARS-CoV-2 in severely ill COVID-19 patients, then longitudinal monitoring of plasma IL-6 levels before and after initiation of steroids may be warranted in clinical practice. Such practice will help determine whether a patient is likely to respond to corticosteroid therapy alone or if they should be considered as candidates for alternative intervention, including combination therapies with IL-6 receptor antagonists like Tocilizumab (Actemra) and Sarilumab (Kevzara), or direct IL-6 antibodies like Siltuximab (Sylvant).

## Methods

### Cohort definition for patients receiving IL-6 measurements and corticosteroids

All patients who tested positive for SARS-CoV-2 (COVID_pos_), as determined by at least one positive PCR test, within the Mayo Clinic Health System and were hospitalized were selected as candidates for further analysis. These COVID_pos_ patients were then filtered by requiring an administration of a systemic corticosteroid at some point during their hospital stay; these corticosteroids included dexamethasone, prednisone, triamcinolone, methylprednisolone, prednisolone, betamethasone, and hydrocortisone. Tocilizumab was also included as a control given it’s known effect on IL-6 signaling. In patients who received a systemic corticosteroid, we required that they underwent plasma IL-6 testing at least once both before and after the first administration of any of the corticosteroid agents listed above. Only 13 patients met these criteria, with 7 patients receiving IL-6 testing before and after methylprednisolone administration, 4 patients receiving IL-6 testing before and after prednisone administration, and 4 patients receiving IL-6 testing before and after hydrocortisone administration. 2 patients overlapped drug categories, with one receiving first administrations of both methylprednisolone and hydrocortisone between IL-6 tests and the other receiving first administrations of both prednisone and hydrocortisone between IL-6 tests). In total, six patients received IL-6 testing both before and after any first corticosteroid administration within the timeframe considered (-25 days to +64 days). In addition to IL-6 levels and corticosteroid/tocilizumab administration data, we extracted outcomes including death, admission to an ICU, and length of time in ICU, as well as demographic information such as age, sex, and race.

For patients with IL-6 measured post-corticosteroid administration, over half of the patients (11 out of 21) were in the ICU at least 15 days or did not survive and had IL-6 levels above 10 pg/mL. As a corollary, fewer patients given corticosteroids (30%, n=21) had high IL-6 levels and shorter ICU stays compared to tocilizumab (80%, n=5), azithromycin (61%, n=49), and hydroxychloroquine (60%, n=10). While not statistically significant due to small cohort sizes, together these results may indicate that patients with high IL-6 levels post-corticosteroids are more likely to have longer ICU durations compared to the other drugs considered here. This points to the necessity for IL-6 measurement post-corticosteroid administration to monitor if further intervention is needed.

### Characterizing the survival curves of hospitalized and ICU COVID-19 patients

Kaplan-Meier curves were generated for 185 COVID-19 positive patients that were hospitalized at the Mayo Clinic (**Figure 8**). Patient cohorts were defined by each drug or drug class that was administered. Patients receiving multiple drugs of different classes were counted as part of both cohorts. The drugs analyzed include corticosteroids, antivirals (remdesivir, lopinavir-ritonavir), antibacterial azithromycin, hydroxychloroquine, and the IL-6 inhibitor tocilizumab. As shown, all 11 hospitalized patients receiving tocilizumab survived 60 days post diagnosis. Hospitalized cohorts receiving corticosteroids (n=95) and azithromycin (n=123) have similar survival rates between 90-95% when considering all hospitalized patients. Hydroxychloroquine (n=21) also has a 60-day post-diagnosis survival rate of 90%, but compared to corticosteroids and azithromycin, mortality occurred much earlier in the hydroxychloroquine cohort, i.e. 2 out of 21 patients died within 5 days of COVID-19 diagnosis. However, when considering more severe COVID patients requiring ICU admission, this subset of hydroxychloroquine patients (n=11) have a 100% survival rate, the azithromycin cohort (n=64) remains relatively unchanged (92% vs. 93% for all hospitalized patients), and the corticosteroid cohort survival rate decreased from 91% to 84% at 60 days post-diagnosis. In both hospitalized and ICU patient cohorts, antivirals performed most poorly, with the caveat that these cohorts also have the smallest patient counts. Similar trends were also observed in the subset of patients with IL-6 measurements in these cohorts.

**Figure 8.**
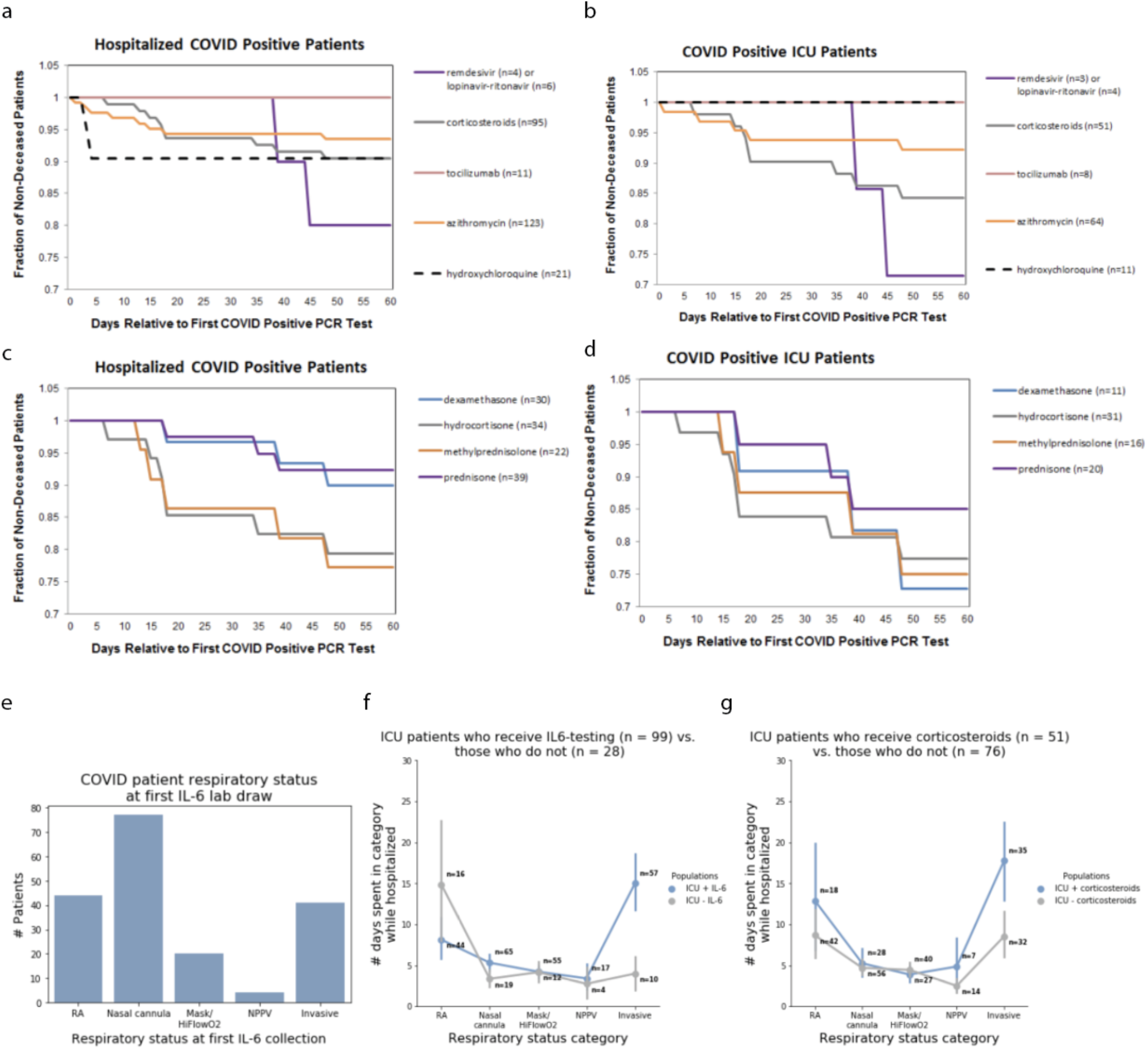
Survival curves and respiratory status for the study cohort. Survival curves for – **(a)** hospitalized and (**b**) ICU -- COVID-19 patients who received corticosteroids, tocilizumab, azithromycin, hydroxychloroquine, or antivirals (remdesivir or lopinavir-ritonavir) following their SARS-CoV-2-positive PCR test. **(c)** Survival curves for each corticosteroid administered patient who is hospitalized, split up by the exact corticosteroid drug. **(d)** Survival curves for each corticosteroid administered patient who is in the ICU, split up by the exact corticosteroid drug, i.e. dexamethasone, hydrocortisone, methylprednisolone, or prednisone. **(e)** Bar plot of COVID-19 patient respiratory status at first IL-6 lab draw, showing that ICU patients who receive IL-6 testing are typically initially tested early in the course of respiratory failure (panel A), **(f)** yet spend a longer time in critical respiratory failure, indicating that physicians may be using IL-6 as part of their assessment of patients who are rapidly progressing towards critical respiratory failure. Here, RA indicates that the patient is breathing room air. Nasal cannula includes all nasal cannula with oxygen flow rates up to 15L/min. Mask/HiFlowO2 includes devices designed to increase the percentage of oxygen inspired with each breath, such as a non-rebreather, Oxymizer, OxyMask, high flow nasal cannula, etc. NPPV includes all forms of non-invasive positive pressure ventilation, such as CPAP (continuous positive airway pressure), BiPAP (bilevel positive airway pressure), and oxygen-supplemented CPAP/BiPAP. Invasive ventilation includes intubation and mechanical ventilation. **(g)** Across every respiratory status, COVID-19 patients who received corticosteroids generally are worse from a respiratory standpoint than patients who did not receive corticosteroids.

To further understand the specific effects of the different corticosteroids given to COVID positive patients, survival rates of hospitalized and ICU patients given dexamethasone, hydrocortisone, prednisone and methylprednisolone were also generated. For hospitalized patients, dexamethasone (n=30) and prednisone (n=39) perform similarly (90% and 92% 60-day survival, respectively) and have higher 60-day survival post-diagnosis compared to hydrocortisone (n=34) and methylprednisolone (n=22) (79% and 77% 60-day survival, respectively). While a decrease in survival is observed in the more severe subset of each of these cohorts who are admitted to the ICU, dexamethasone (n=11) shows the most significant decrease in 60-day survival, dropping from 90% to 72%.

### Comparison of patient IL6/ICU pattern between drug classes

We defined 4 categories of patient IL-6 level/ICU stay length (i.e. the 4 quadrants mentioned in **Figure 1b**). We split patient IL-6 level into 2 categories (high and low) and ICU stay length into 2 categories (*poor outcome* -- i.e. 15+ days or ending in death, and *not poor outcome* - - i.e. <15 days in ICU). We then compared the proportion of high IL-6 patients who had poor outcomes for (i) cohort of patients who took corticosteroids vs (ii) the cohort of patients who did not take corticosteroids (but did take one of our other drugs of interest); as well as the proportion of low IL-6 patients who had poor outcomes in each of those cohorts. We performed a Fisher exact test to compute p-values on these proportion comparisons.

This research was conducted under IRB 20-003278, “Study of COVID-19 patient characteristics with augmented curation of Electronic Health Records (EHR) to inform strategic and operational decisions”. All analysis of EHRs was performed in the privacy-preserving environment secured and controlled by the Mayo Clinic. nference and the Mayo Clinic subscribe to the basic ethical principles underlying the conduct of research involving human subjects as set forth in the Belmont Report and strictly ensures compliance with the Common Rule in the Code of Federal Regulations (45 CFR 46) on Protection of Human Subjects.

### Bulk RNA-Sequencing Analysis

#### Data accession and processing

Datasets were downloaded in raw fastq format and uniformly processed using *salmon* as previously described^14^.

#### Global expression analysis

To identify highly expressing cells and tissues for a given gene, the following steps are followed:

1. Plot the distribution of gene expression (in units of transcripts per million, or TPM) across all samples from all studies.
2. Divide distribution into “High Expression Group” (e.g., cells in top 5% of expressing samples for query gene) and “Low Expression Group” (e.g. cells in bottom 25% of expressing samples for query gene).
3. Count the number of individual samples from each annotated cell or tissue type falling in the High and Low Expression Groups. Note that cells and tissues are extracted from sample-level metadata available through the Gene Expression Omnibus (GEO) and other databases including the Genotype-Tissue Expression project (GTEx), The Cancer Genome Atlas (TCGA), and the Cancer Cell Line Encyclopedia (CCLE).
4. Compute Fisher’s Exact Test p-value to measure the enrichment of cell type C (or tissue T) among the High Expression Group. Enrichment Scores displayed correspond to - log_10_(adjusted p-value), where p-values are adjusted using the Benjamini-Hochberg (BH) correction.

### Single Cell RNA-Sequencing Analysis

#### Data accession and processing

Datasets were downloaded and processed as previously described^14^, and processed datasets have been made available for investigation upon registration in the nferX Single Cell platform (https://academia.nferx.com/).

#### Global expression analysis

To identify highly expressing cells for a given gene, the following steps are followed:

1. Plot the distribution of gene expression (in units of counts per 10,000, or CP10K) across all single cells from all studies.
2. Divide distribution into “High Expression Group” (e.g., cells in top 10% of expressing samples for query gene) and “Low Expression Group” (e.g. cells in bottom 90% of expressing samples for query gene).
3. Count the number of individual cells from each annotated cell population (or tissue) falling in the High and Low Expression Groups.
4. Compute Fisher’s Exact Test p-value to measure the enrichment of cell population C (or tissue T) among the High Expression Group. Enrichment Scores displayed correspond to - log_10_(adjusted p-value), where p-values are adjusted using the Benjamini-Hochberg (BH) correction.

#### Coexpression analysis

For a given set of genes, a single coexpression vector is computed as the geometric mean of CP10K values of all genes in each cell. The geometric mean is used as a coexpression metric as it will only yield a positive value in cells which express all genes in the defined set (i.e. one or more zero values in an individual cell will result in a coexpression value of zero for that cell). As such, all cells with a coexpression value (CP10K_gm_) greater than zero can be considered as “coexpressing cells”, whereas all cells with a CP10K_gm_ values equal to zero can be considered as “non-coexpressing cells.” After this coexpression vector has been computed, it is treated identically to a gene expression vector for a single gene in the context of the Global Expression or single study-level analyses described above.

### Institutional Review Board (IRB) approval for this research

This research was conducted under IRB 20-003278, “Study of COVID-19 patient characteristics with augmented curation of Electronic Health Records (EHR) to inform strategic and operational decisions”. All analysis of EHRs was performed in the privacy-preserving environment secured and controlled by the Mayo Clinic. nference and the Mayo Clinic subscribe to the basic ethical principles underlying the conduct of research involving human subjects as set forth in the Belmont Report and strictly ensures compliance with the Common Rule in the Code of Federal Regulations (45 CFR 46) on Protection of Human Subjects. For more information, please visit https://www.mayo.edu/research/institutional-review-board/overview

## Data Availability

The data analyzed in this manuscript was accessed under IRB 20-003278 - Study of COVID-19 patient characteristics with augmented curation of Electronic Health Records (EHR) to inform strategic and operational decisions. All analysis was performed in a privacy-preserving environment. For more information, please visit https://www.mayo.edu/research/institutional-review-board/overview

## Acknowledgements

The authors thank Patrick Lenehan, Murali Aravamudan, Travis Hughes, Andrew Danielsen, Vineet Agarwal, Sankar Ardhanari, Jason Ross, Najat Khan, Anuli Anyanwu Ofili, Kristopher Standish, and James Merson for their helpful feedback on this manuscript.

## Supplementary Material

**Figure 1 - Supplementary Figure 1.**
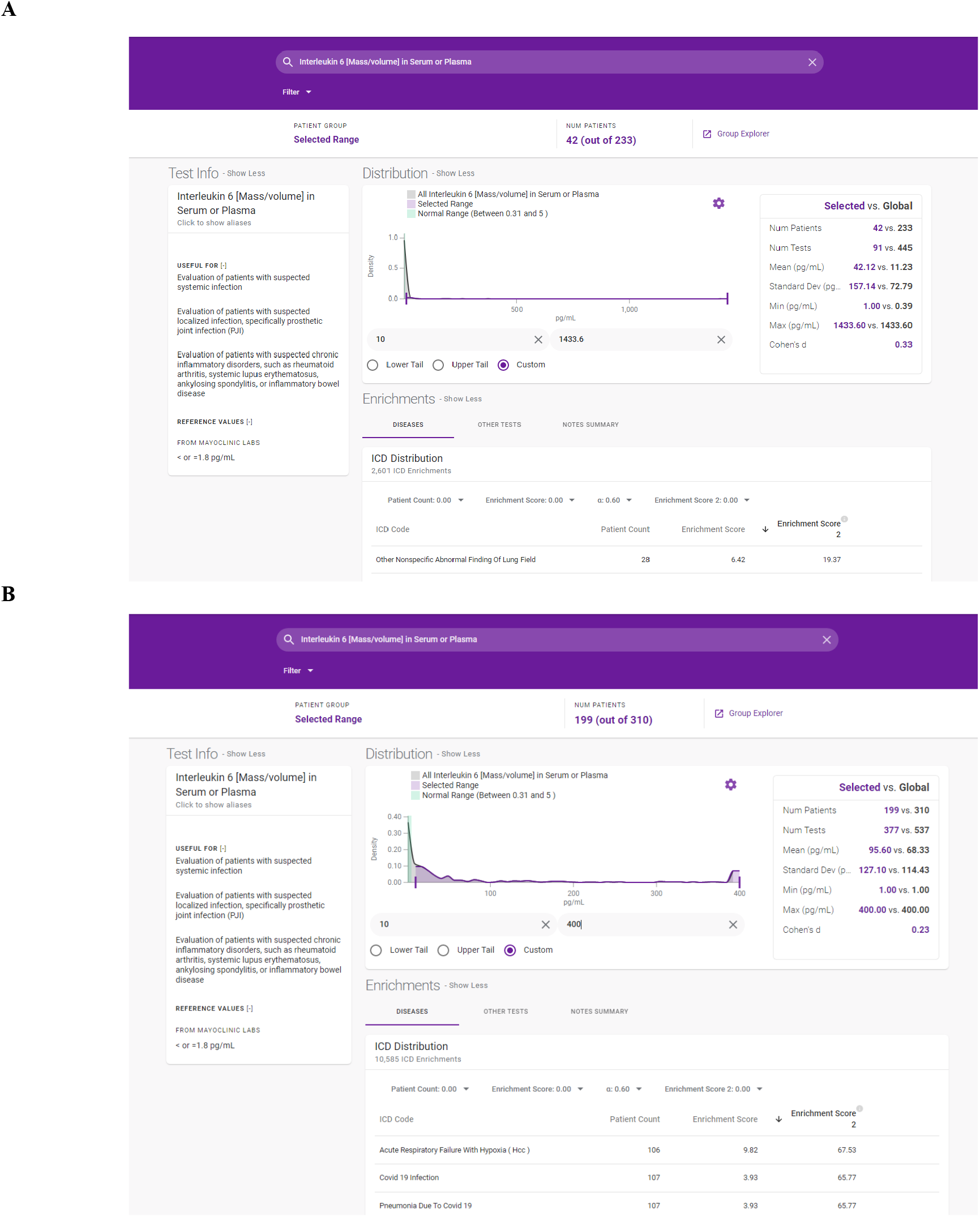
Distribution of serum or plasma IL-6 measurements for a subset of 700k patients at the Mayo Clinic taken in 2019 **(A)** and 2020 **(B).** Also shown are the enriched ICD codes for the patients in the right tail of the distribution with abnormally high IL-6 levels.

**Figure 1 - Supplementary Figure 2.**
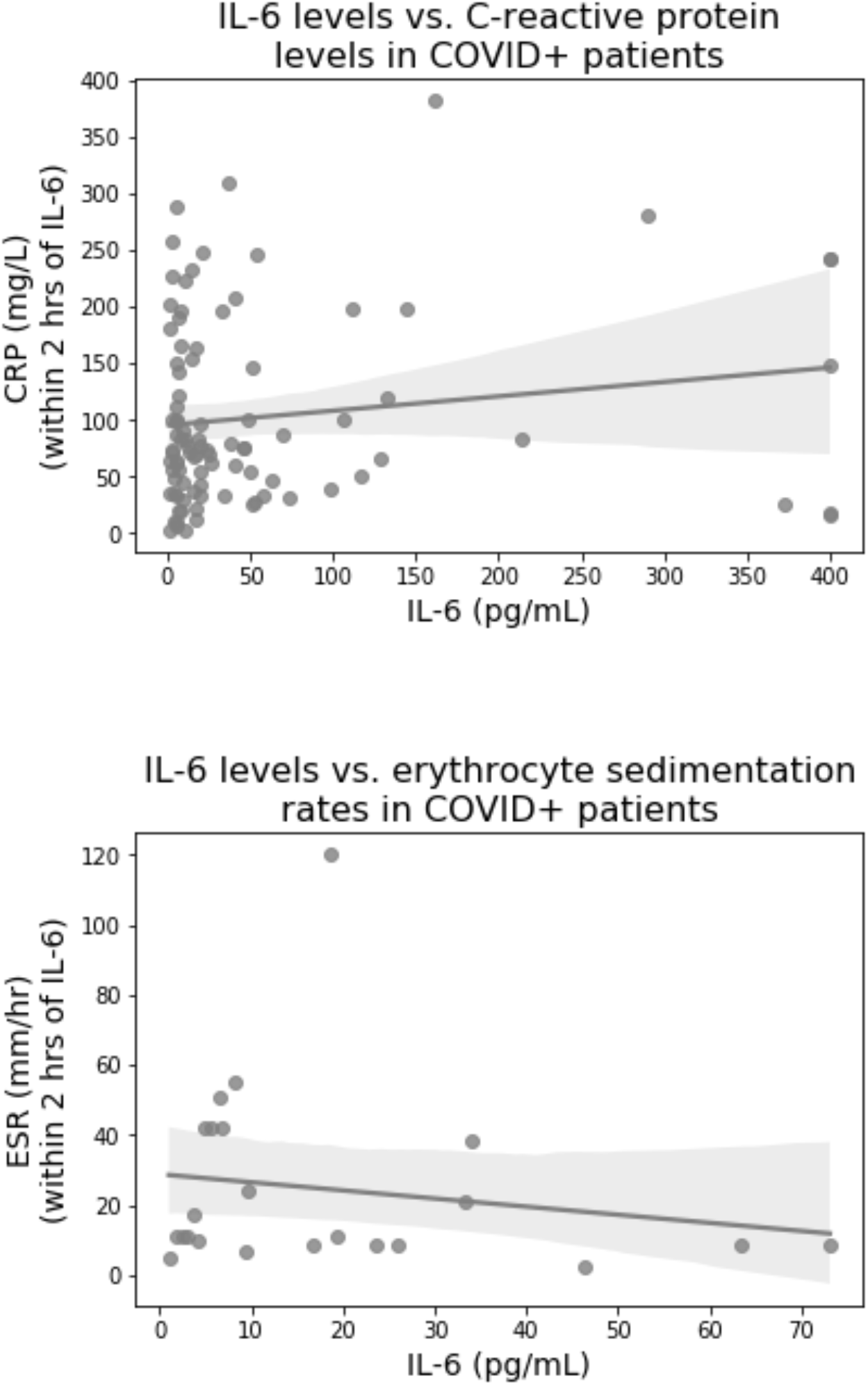
Relationship between the inflammatory markers C-reactive protein (CRP) and the erythrocyte sedimentation rate and IL-6 in hospitalized COVID-19 patients. In each case, blood for inflammatory marker measurement was collected within 2 hrs of IL-6 blood collection. The linear regression is shown with a dark grey line, and 95% confidence intervals around the regression are shown in light grey. In both cases, no significant relationship between IL-6 and levels of the inflammatory marker is found.

**Figure 1 - Supplementary Figure 3.**
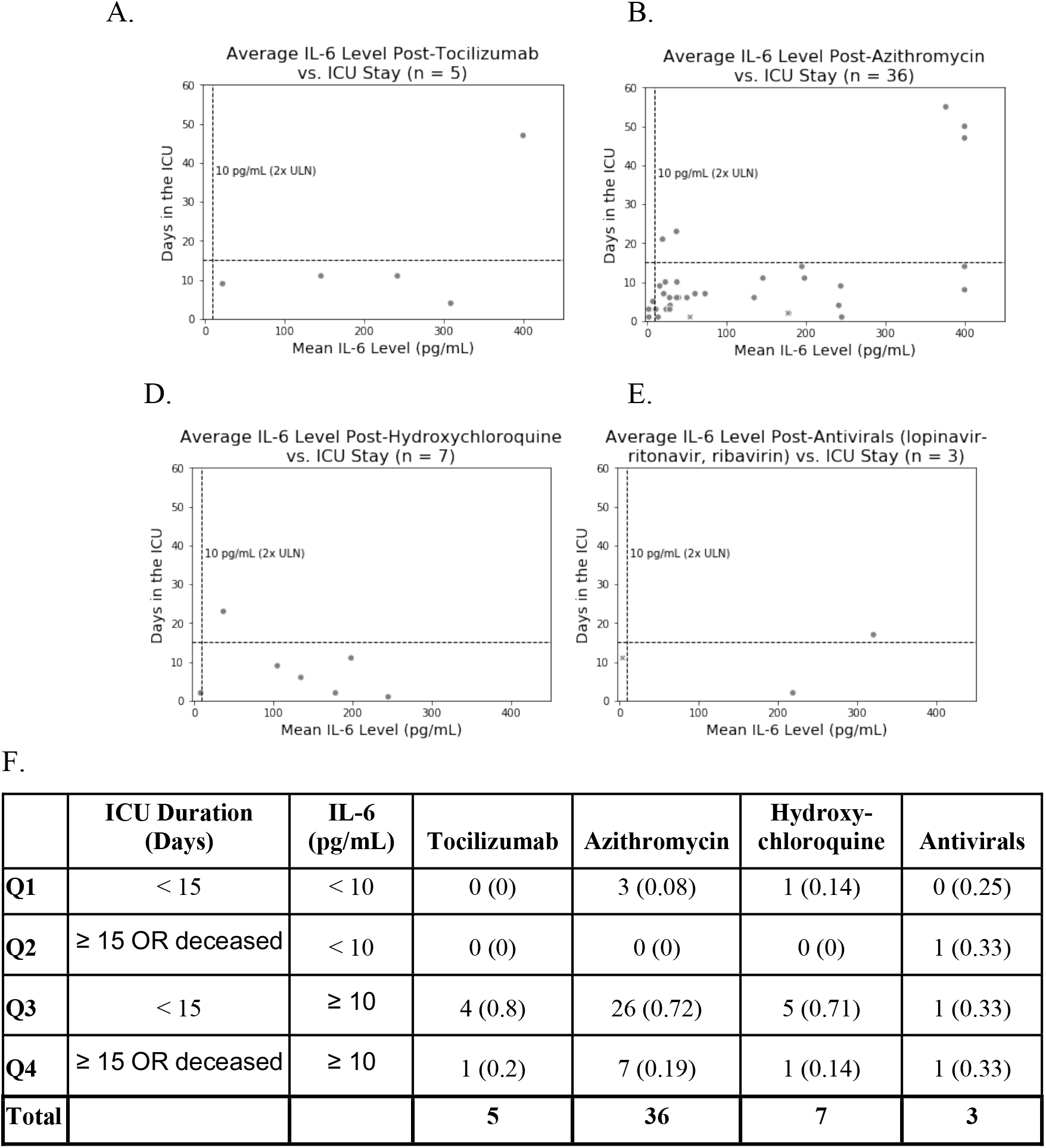
Relationship between IL-6 levels and ICU duration for COVID-19 ICU patients who had at least one plasma IL-6 measurement after therapeutic administration. Longitudinal measurements of plasma IL-6 in patients after they received (**A)** Non-topical corticosteroids (**B**) Tocilizumab (**C**) Azithromycin (**D**) Hydroxychloroquine (**E**) antivirals, at least once at or after the diagnosis of COVID-19 via SARS-CoV-2 PCR. (**F**) Patient counts and proportions for each quadrant, where deceased patients were counted with those in the ICU ≥ 15 days.

**Figure 2 - Supplementary Figure 1.**
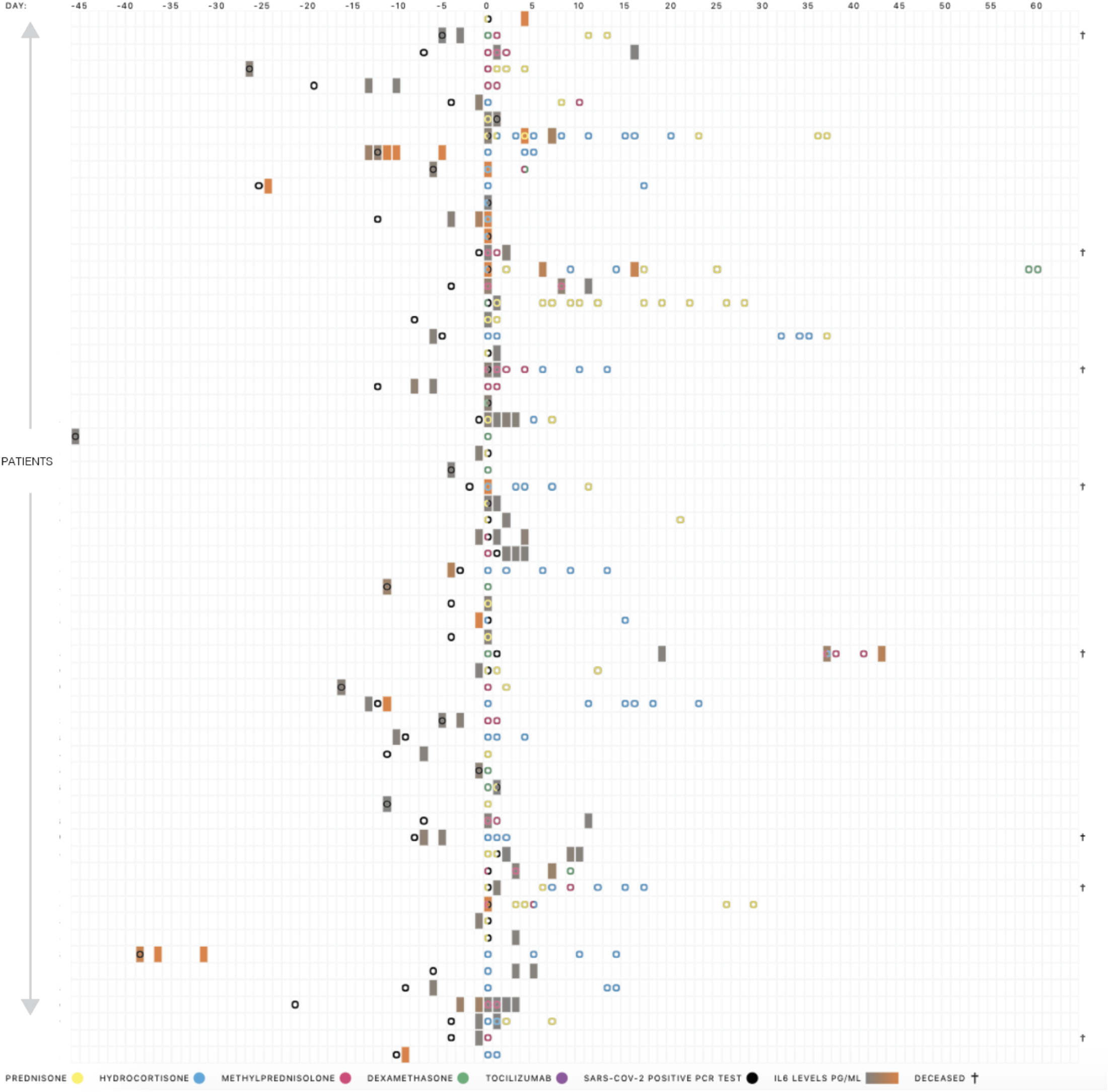
Characterization of the overall cohort of n = 63 COVID_pos_ patients in the ICU that received a SARS-CoV-2 positive PCR test (black), a therapeutic treatment (other colors) and plasma IL-6 lab test (gray-to-orange gradient).

**Figure 3 - Supplementary Figure 1.**
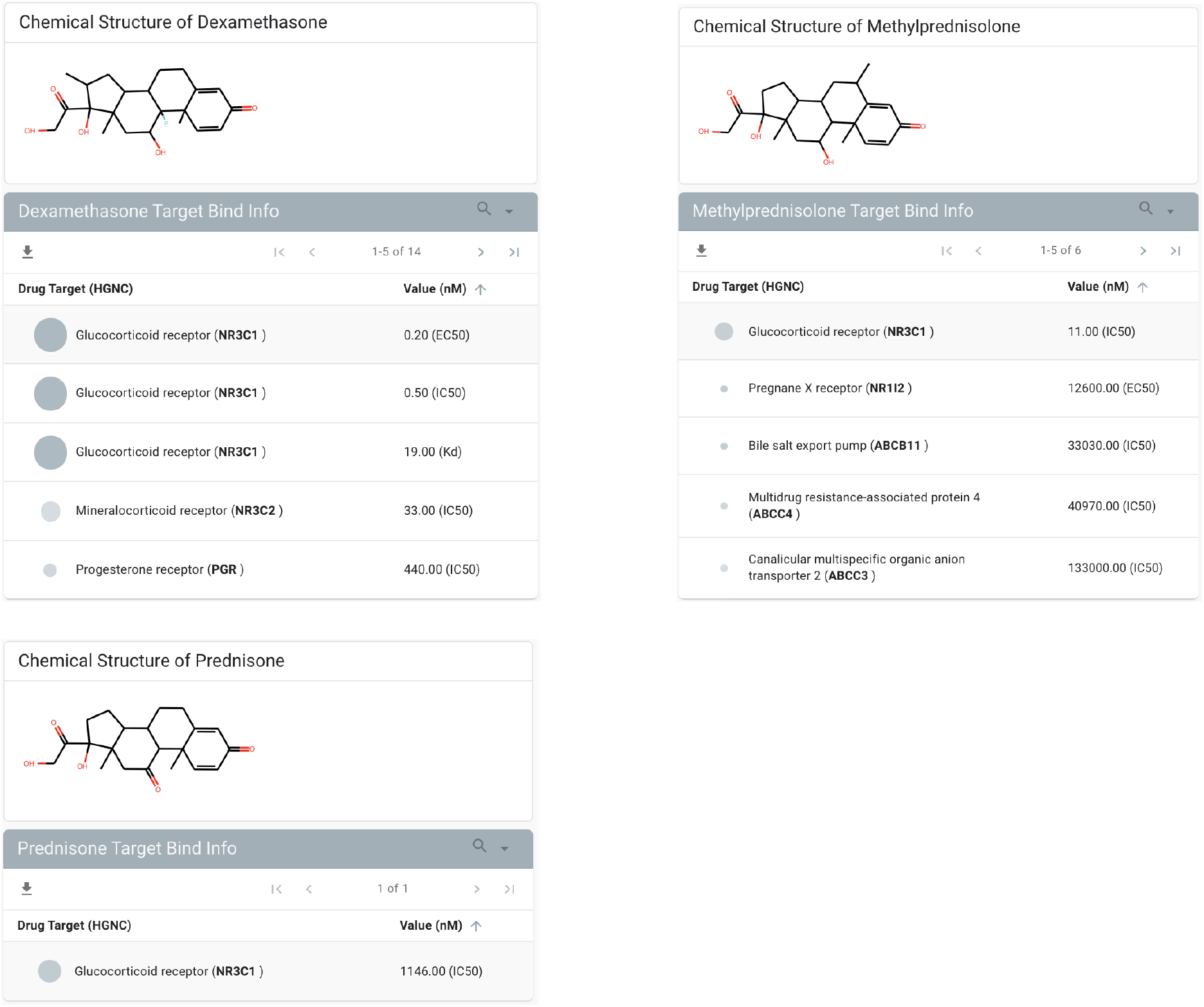
Characterization of molecular targets of dexamethasone, methylprednisolone, and prednisone. NR3C1 is the highest affinity target for each of these corticosteroids.

**Figure 3 - Supplementary Figure 2.**
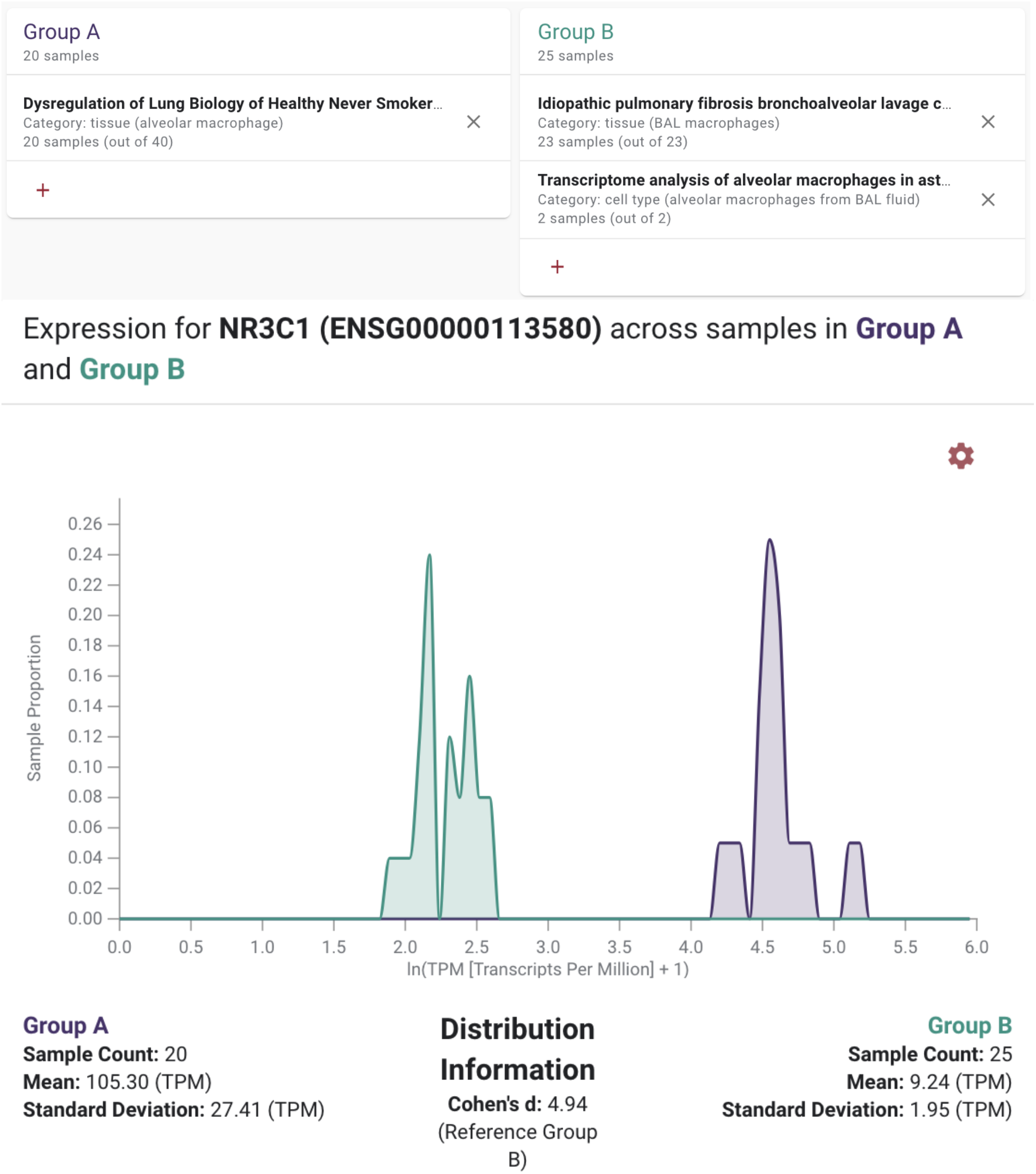
Expression of NR3C1 by bulk RNA-sequencing in alveolar macrophages from three independent studies. In one study (Group A, purple), alveolar macrophages were found to express high levels of NR3C1, with 13 of 20 samples falling in the top 5% of NR3C1 expression considering all human samples deposited in GEO. In two other studies (Group B, green), alveolar macrophages expressed lower but still appreciable levels of NR3C1.

**Figure 3 - Supplementary Figure 3.**
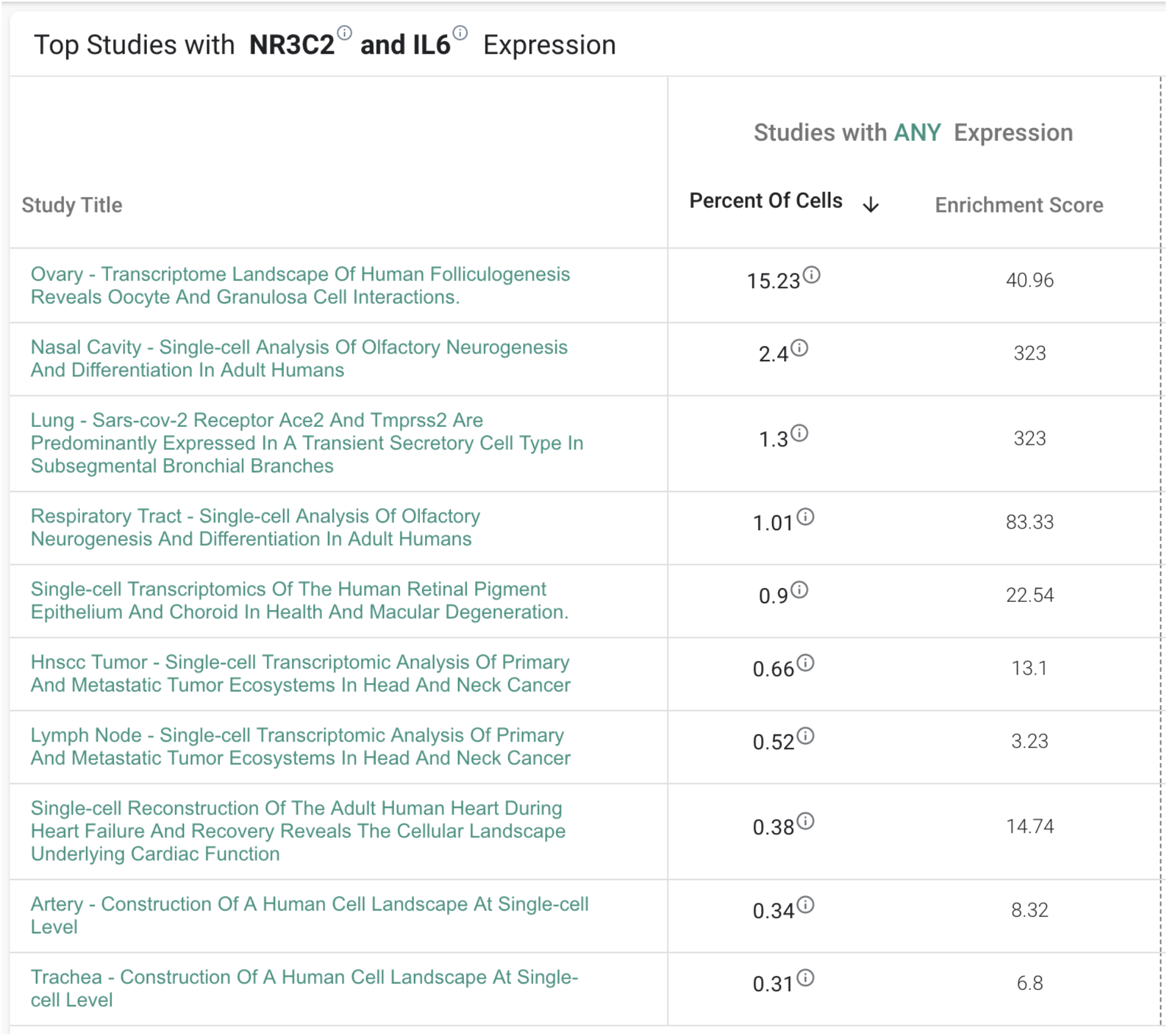
Single cell RNA-seq studies showing non-zero (any) co-expression of NR3C2 and IL-6. Primary oocytes, granulocytes, and secondary oocytes of the ovaries show significant co-expression of NR3C2 and IL-6. The other cell types only have minor populations of cells that co-express both NR3C2 and IL-6.

